# The association between plasma chemokines and breast cancer risk and prognosis: A Mendelian randomization study

**DOI:** 10.1101/2022.03.04.22271238

**Authors:** Xingxing Yu, Yanyu Zhang, Yuxiang Lin, Shuqing Zou, Pingxiu Zhu, Mengjie Song, Fangmeng Fu, Haomin Yang

## Abstract

**Background:** Despite the potential role of several chemokines in the migration of cytotoxic immune cells to prohibit breast cancer cell proliferation, a comprehensive view of chemokines and the risk and prognosis of breast cancer is scarce, and little is known about their causal associations.

**Methods:** With a two-sample Mendelian randomization (MR) approach, genetic instruments associated with 30 plasma chemokines were created. Their genetic associations with breast cancer risk and survival were extracted from the recent genome-wide association study, with available survival information for 96,661 patients. We further tested the associations between the polygenetic risk score (PRS) for chemokines and breast cancer in the UK Biobank cohort using logistic regression models. The association between chemokine expression in tumors and breast cancer survival was analyzed in the TCGA cohort with Cox regression models.

**Results:** Plasma CCL5 was causally associated with the risk of breast cancer in the MR analysis, which was significant in the luminal and HER-2 enriched subtypes and further confirmed using PRS analysis (OR=0.94, 95% CI=0.89-1.00). A potential causal association with breast cancer survival was only found for plasma CCL19, especially for ER-positive patients. In addition, we also found an inverse association between CCL19 expression in tumors and breast cancer overall and relapse-free survival (HR=0.58, 95% CI=0.35-0.95).

**Conclusion:** We observed an inverse association between genetic predisposition to CCL5 and the risk of breast cancer, while CCL19 was associated with breast cancer survival. These associations suggested the potential of these chemokines as tools for breast cancer prevention and treatment.

## Introduction

Breast cancer is one of the most common malignant tumors, which seriously affects the physical and mental health of women worldwide and accounts for 24.2% and 15.0% of the total number of new cancers and deaths in women, respectively [1]. Although breast cancer has caused a considerable global burden, its etiology is not yet completely understood. Currently, the known breast cancer risk factors include age, sex, race, family history, genetic mutations, early menarche/late menopause and hormone replacement therapy, while evidence for the causal effect of inflammation in breast cancer carcinogenesis is still limited [2, 3]. To prevent the incidence of breast cancer and find potential therapeutic targets, we need to better understand the role of inflammatory biomarkers in breast cancer.

At the first stage of malignant evolution, chronic inflammation may play an important role during the initiation, development and prognosis of breast cancer [4], in which chemokines might regulate the migration of immune cells toward the tumor and act as biomarkers for breast cancer. Studies have shown that CCL-4, -7, -8, -11, -15, -16, -19, -22, -23, -24, -25 and CXCL-5, -8, -9, and -16 were increased in breast cancer compared with normal adjacent breast tissue [5], and genetic polymorphisms near genes encoding CCL-2, -4, -5, -20, -22, CXCL-10, -12 and XCL-1 were found to be associated with the risk and prognosis of breast cancer [6-13]. However, traditional observational studies are susceptible to reverse causation and residual confounding [14]. Contrary to previous studies, Bowen Chen et al [15] used several bioinformatics analysis tools to analyze the expression data of CC chemokines in patients with BC and yielded inconsistent results. Therefore, it is still a challenge to fully understand the causal association between chemokines and breast cancer.

Mendelian randomization (MR) uses genetic variation as an instrumental variable (IV) to infer the association between exposure factors and disease outcomes. Because the genotypes are presumed to be randomly allocated in the process of gamete formation, MR analyses are largely free of reverse causality and less sensitive to confounding factors, which overcomes the limitations of conventional observational studies [14, 16].

In the present study, we implemented a two-sample MR design to investigate the potential causal effect of plasma chemokines on the risk and prognosis of breast cancer and to analyze their molecular subtypes to provide more credible evidence on the potential role of chemokines in the development of breast cancer. We further used the UK Biobank and TCGA cohorts to validate our findings.

## Methods

### Study population

Mendelian randomization analysis was used to assess the causal association between plasma chemokines and breast cancer risk. In the first step, the associations were tested using a two-sample approach with summary statistics from genome-wide association studies (GWAS), which does not need individual-level data[17]. Genetic instrumental variables for plasma chemokines were created by identifying single nucleotide polymorphisms (SNPs) associated with 41 plasma chemokines in a recent GWAS on the human plasma proteome at the GWAS significant level (*p*<5×10^−8^) [18]. From the 5965 SNPs reported in the original study, 84 SNPs for 30 plasma chemokines were selected after excluding SNPs with linkage disequilibrium (LD threshold: *r*^2^<0.01) and palindromes. The SNPs used in this study and their corresponding plasma chemokines and proportion of variance explained are listed in sTable 1. We used R^2^ and F statistics to estimate the proportion of variance explained by SNPs, with the following formulas 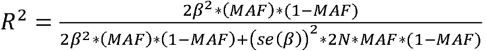. and 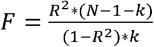, in which is the coefficient for the SNP, MAF is the minor allele frequency, *se*(β) is the standard error of the coefficient, N is the sample size and k is the number of SNPs used.

GWAS summary statistics for breast cancer risk overall and by molecular subtypes (luminal A, luminal B, Her-2 enriched and triple negative) were downloaded from the Breast Cancer Association Consortium [19], with 133,384 breast cancer cases and 113,789 controls. The GWAS summary statistics for breast cancer survival were also downloaded from BCAC, including 96,661 patients with 7,697 breast cancer mortality cases; among them, 64,171 were ER-positive patients, and 16,172 were ER-negative [20]. All studies involved in the GWAS on the human plasma proteome and the Breast Cancer Association Consortium were approved by the relevant research ethics committee, and all participants provided informed consent.

For those identified plasma chemokines in the two sample Mendelian randomization studies for breast cancer risk, a single-sample Mendelian randomization was further applied to validate the findings in UK Biobank. Briefly, the UK Biobank is a prospective cohort comprising approximately 502,505 participants (55% women) of various ethnicities, aged 40-69 at recruitment between 2006 and 2010. At enrolment, information on the participants’ health-related factors and socio-demographic characteristics and biological samples were collected in 22 assessment centers around UK.

Blood samples from women in the UK Biobank were genotyped using a custom UK Biobank Axiom Array, comprising 825,927 SNPs. Details of the array design, sample handling, quality control processes and imputation of non-genotyped variants are described elsewhere [21]. Breast cancer cases were obtained through linkage with the national cancer registries in the UK and updated until 2015. The ICD-10 code C50 and ICD-9 code 174 were used to identify breast cancer cases, while the controls were women without a diagnosis of breast cancer. To assess whether breast cancer is associated with a genetic predisposition to plasma chemokines, we selected the same genome-wide significant SNPs used in the two-sample MR. For all individuals, a weighted polygenic risk score (PRS) was calculated using the following formula:

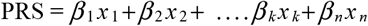

where *β* is the per-allele log odds ratio (OR) of the breast cancer-associated risk allele for SNP _*k*_, *x* _*k*_ is the number of alleles for the same SNP (0, 1, 2), and _*n*_ is the total number of breast cancer SNPs included in the profile. For the analyses, PRS was categorized into tertiles.

To further explore the effect of the identified chemokines associated with breast cancer survival, 1077 women in the TCGA breast cancer cohort were followed for their overall and breast cancer survival. Gene expression data of the chemokines in tumor tissues were also downloaded and categorized into tertiles.

### Statistical Analysis

Several methods were used to estimate the potential causal effect of plasma chemokines on breast cancer risk and prognosis. The main analysis was based on a fixed-effects meta-analysis using an inverse-variance weighting model (IVW), in which β and *se* were calculated using the following formulas 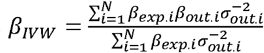 and 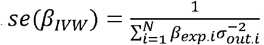 : N is the total number of SNPs, *βexp*.*i* is the coefficient of the *i*-th SNP on the exposure, *βout*.*i* is the coefficient of the *i*-th SNP on the outcome, and *σout*.*i* is the standard error of *βout*.*i*. If only one SNP was identified for the chemokine, the Wald ratio method was used by calculating the ratio of *βexp* and *βout*, while *se* could be approximately estimated using the Delta method. [22] We further performed sensitivity analysis to test horizontal pleiotropy using weighted median and Egger regression methods. The weighted median estimator is consistent with the IVW estimation even when up to 50% of the SNPs are invalid instrumental variables, while MR–Egger regression simply adds an intercept term of pleiotropy into the IVW regression model [23].

To test the key assumption of no horizontal pleiotropy in MR, we further analyzed the effect of plasma chemokines on potential risk factors for breast cancer, e.g., age at menarche, age at menopause, age at smoking initiation, and BMI, using their corresponding GWAS summary statistics [24-26].

To validate the discovered potential causal association between plasma chemokines and breast cancer, we used logistic regression models to calculate the ORs of breast cancer among UK biobank women by tertiles of PRS for plasma chemokines. These associations were adjusted for age, the first ten principal components, and the assessment center. We also used the Cox model to estimate the hazard ratios of overall survival and relapse-free survival in the TCGA cohort by tertiles of gene expression of the chemokines in tumor tissues and adjusted for age at diagnosis, estrogen, progestogen, and tumor stage.

All statistical tests were two-sided and performed using R 3.6.3 and Stata 15.1.

## Results

In the main analysis, the selected SNPs explained 0.9%–36.3% of the variance in plasma chemokines, and the corresponding F-statistic ranged from 29.7–1879.3 (sTable 1), suggesting no evidence of weak instrument bias. A higher plasma CCL5 was associated with a lower risk of breast cancer (OR=0.89, 95% CI: 0.85 to 0.94, p<0.001), which was primarily attributed to the luminal A and luminal B subtypes (OR for luminal A=0.88, 95% CI: 0.82 to 0.94; OR for luminal B=0.80, 95% CI: 0.69 to 0.94). Plasma growth-regulated alpha protein and Ck-beta-8 were also associated with the risk of breast cancer, although the p value did not survive the Bonferroni correction for multiple testing (Figure 1, sTable 2).

**Figure 1.**
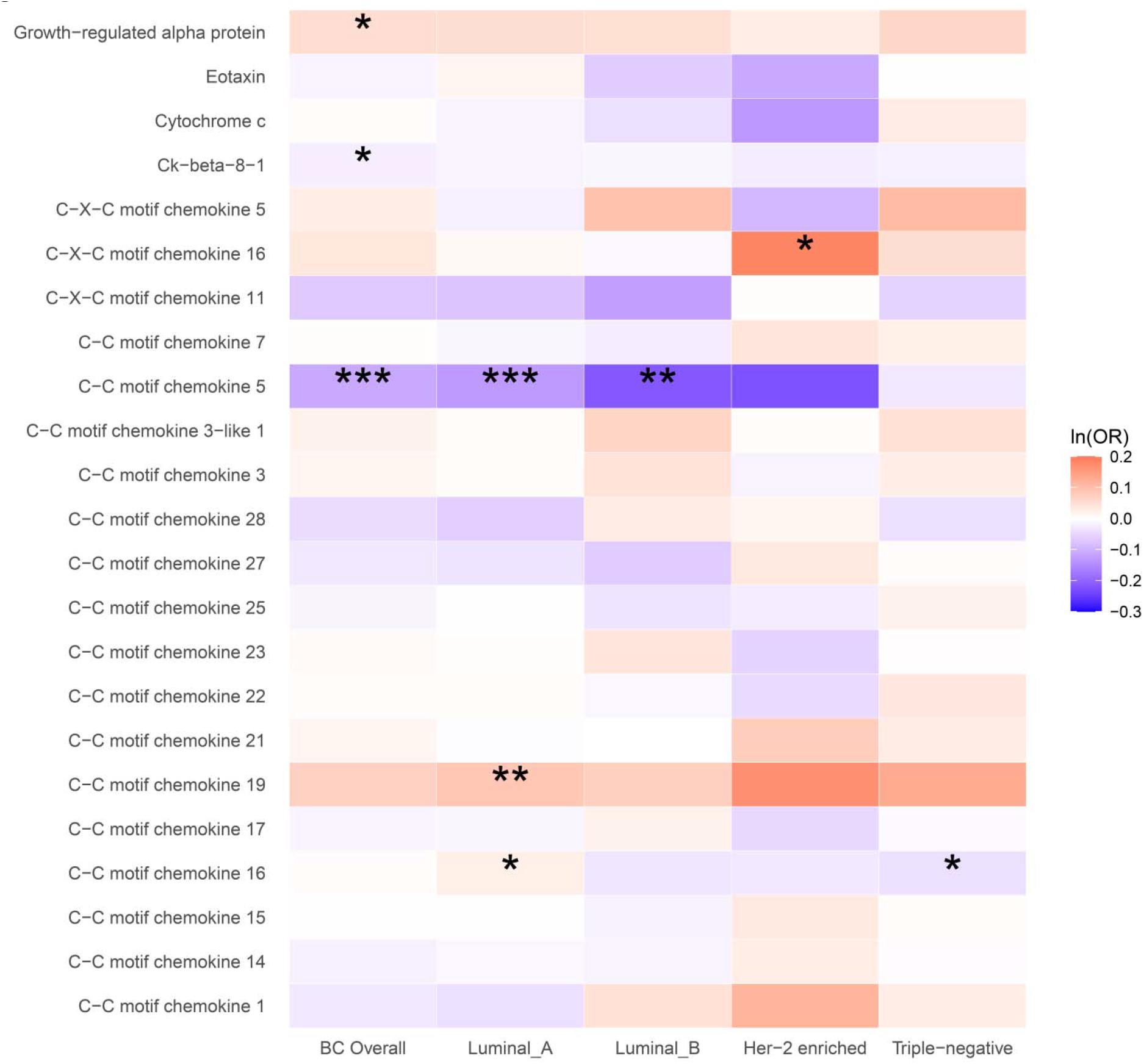
The causal association between plasma chemokines and breast cancer risk overall and by molecular subtype. OR: odds ratio. Statistical significance is indicated by *** (p<0.001), ** (p<0.01) and * (p<0.05). The estimates are based on the fixed effect of the inverse variance weighted method or Wald ratio method for meta-analyzing individual SNP results. ORs show the risk of breast cancer overall and by subtypes per SD increase of ranked-inverse normalized concentration in exposure to plasma chemokines.

In the subtype-specific analysis, plasma CCL19 was associated with luminal A breast cancer, while CXCL16 was associated with HER-2-enriched breast cancer. In addition, plasma CCL16 was positively associated with the risk of luminal A breast cancer but inversely associated with triple-negative breast cancer. Sensitivity analysis using MR Egger and weighted median methods yielded similar results (sTable 2).

To validate the findings in the two-sample MR analysis, 264,187 women who had available genotyped data from UK Biobank were involved in the analysis, with 14760 breast cancer patients. In multivariable logistic regression analysis, a higher polygenic risk scores for plasma CCL5 was inversely associated with breast cancer (OR for the highest tertile =0.94, 95% CI=0.89-1.00, *P*=0.045). However, no statistically significant associations for plasma growth-regulated alpha protein, Ck-beta-8 and breast cancer were found. (Table 1).

**Table 1.** The association between polygenic risk scores for chemokines and breast cancer in the UK Biobank

| Chemokine | No. | No. of cases | OR (95% CI) | *P* value |
| --- | --- | --- | --- | --- |
| Growth-regulated alpha protein |  |  |  |  |
| Tertile 1 | 131908 | 7461 | 1.00(REF) |  |
| Tertile 2 | 91150 | 5184 | 1.02 (0.98-1.06) | 0.251 |
| Tertile 3 | 41129 | 2115 | 0.97 (0.92-1.02) | 0.194 |
| C-C motif chemokine 5 |  |  |  |  |
| Tertile 1 | 136038 | 7636 | 1.00(REF) |  |
| Tertile 2 | 98239 | 5503 | 0.98 (0.95-1.02) | 0.273 |
| Tertile 3 | 29910 | 1621 | **0.94 (0.89-1.00)** | 0.045 |
| Ck-beta-8-1 |  |  |  |  |
| Tertile 1 | 91937 | 5073 | 1.00(REF) |  |
| Tertile 2 | 121826 | 6799 | 0.97 (0.93-1.00) | 0.073 |
| Tertile 3 | 50424 | 2888 | 0.99 (0.94-1.04) | 0.664 |

The model was adjusted for age at recruitment, assessment center and top 10 principal genetic components.

In the analysis of breast cancer survival, we found a potential causal effect of plasma CCL19 on breast cancer survival (HR=0.85, 95% CI=0.75-0.96, *P*<0.01), especially for ER-positive breast cancer patients (HR=0.82, 95% CI=0.70-0.97) (Figure 2, sTable 3). Moreover, in the TCGA breast cancer patient cohort, CCL19 expression in tumor tissues was associated with increased overall and relapse-free survival (HR for the last tertile vs. first tertile=0.58, 95% CI=0.35-0.95). The association was also validated for ER-positive patients (Table 2).

**Figure 2.**
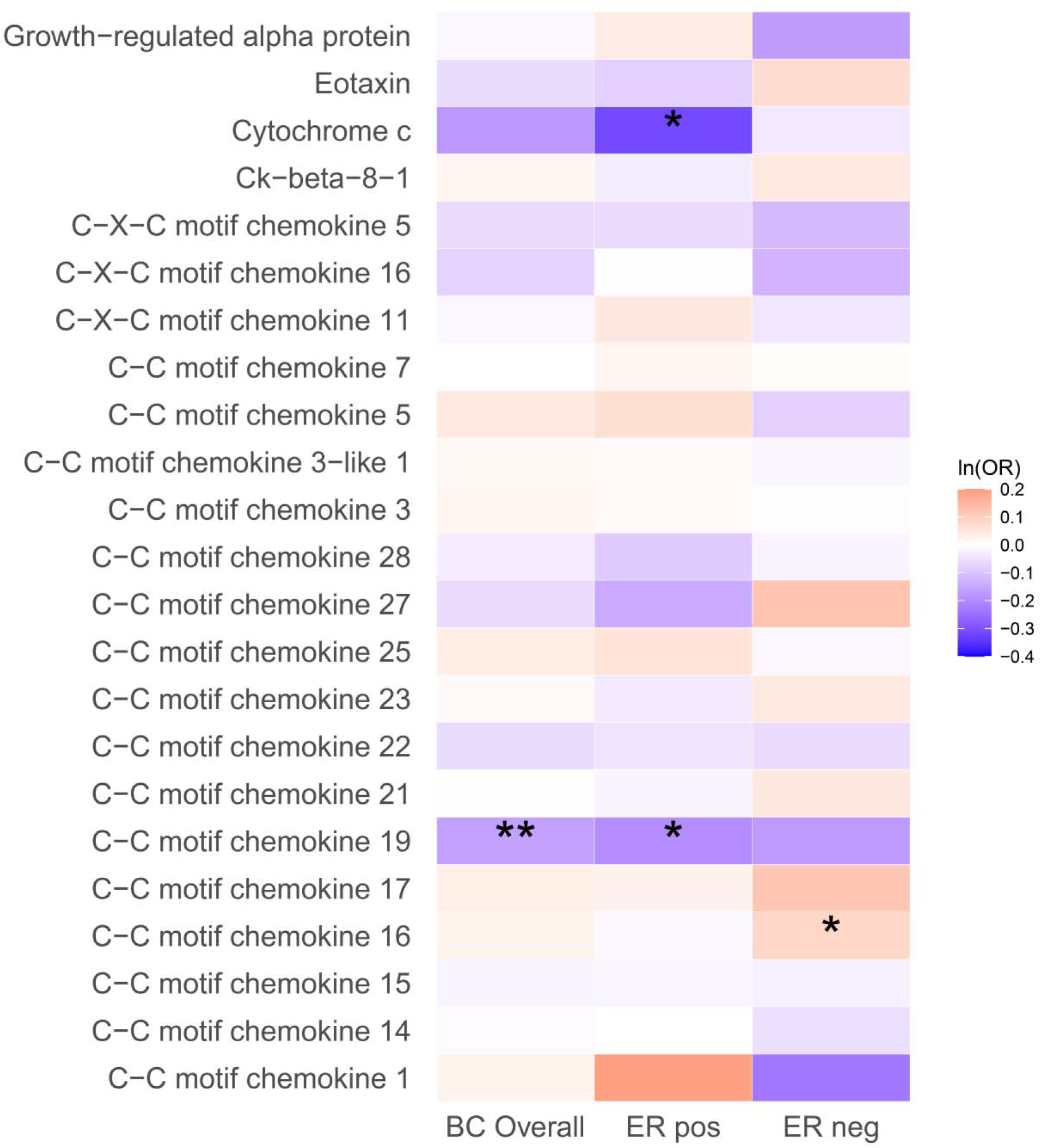
The causal association between plasma chemokines and breast cancer survival. HR: hazard ratio. Statistical significance is indicated by ** (p<0.01) and * (p<0.05). The estimates are based on the fixed effect of the inverse variance weighted method or Wald ratio method for meta-analyzing individual SNP results. HRs show the risk of breast cancer overall and by subtypes per SD increase of ranked-inverse normalized concentration in exposure to plasma chemokines.

**Table 2.** The association between CCL19 and survival of breast cancer patients in TCGA cohort.

|  | Total No. | Overall survival |  | Relapse free survival |  |
| --- | --- | --- | --- | --- | --- |
|  |  | No. of cases | HR (95% CI) | No. of cases | HR (95% CI) |
| Breast cancer overall |  |  |  |  |  |
| Tertile 1 | 358 | 64 | 1.00(REF) | 39 | 1.00(REF) |
| Tertile 2 | 358 | 46 | **0.64 (0.43-0.96)** | 23 | **0.49 (0.29-0.85)** |
| Tertile 3 | 357 | 43 | **0.66 (0.43-1.00)** | 34 | **0.58 (0.35-0.95)** |
| ER+ breast cancer |  |  |  |  |  |
| Tertile 1 | 264 | 46 | 1.00(REF) | 26 | 1.00(REF) |
| Tertile 2 | 275 | 33 | **0.52 (0.32-0.84)** | 19 | **0.51 (0.27-0.97)** |
| Tertile 3 | 249 | 22 | **0.44 (0.26-0.77)** | 17 | **0.43 (0.22-0.82)** |
| ER- breast cancer |  |  |  |  |  |
| Tertile 1 | 72 | 12 | 1.00(REF) | 10 | 1.00(REF) |
| Tertile 2 | 72 | 10 | 0.92 (0.38-2.24) | 4 | 0.40 (0.12-1.38) |
| Tertile 3 | 94 | 20 | 1.09 (0.49-2.44) | 16 | 0.81 (0.33-1.98) |

The model was adjusted for age at diagnosis, ER, PR and stage.

MR estimates for the effects of plasma chemokines on breast cancer risk factors indicated little evidence of horizontal pleiotropy (sTable 4).

## Discussion

In this Mendelian randomization study of 30 plasma chemokines, we found an inverse association between genetically predicted plasma CCL5 and breast cancer risk in both the two-sample and one-sample analyses. This association was more pronounced in luminal and HER-2-enriched breast cancer patients. An effect on cancer survival was only found for plasma CCL19, especially for ER-positive patients. In addition, we also found an inverse association between CCL19 expression and breast cancer mortality using tumor tissue samples.

As key proteins in immune cell migration, chemokines have been studied for their effect on inflammation-associated cancers. One previous Mendelian randomization analysis on cytokines found an association between genetically predicted CCL2, CCL4, GRO*α* and the risk of breast cancer. As the SNPs associated with plasma CCL2 and CCL4 did not exceed the GWAS significance threshold, we were not able to replicate these findings in our study. However, both our study and a previous study confirmed the association between GRO*α* and the risk of breast cancer, confirming the role of GRO*α* in breast cancer carcinogenesis and probably from the effect of Th17 cells [27].

Our analysis found an inverse association between CCL5 and breast cancer risk, with a similar effect size in a recent phenome-wide Mendelian randomization [28]. We further suggested that the association was driven by luminal and HER-2-enriched subtypes of breast cancer. Although tumor-derived CCL5 in breast cancer patients was found to be associated with poor prognosis [29], plasma CCL5 might still play an antitumor role, as the function of the CCL5/CCR5 axis varies in different conditions[30]. CCL5 can mediate the migration of immune cells and promote lymphocyte migration in breast cancer cells [31]. Many autoimmune diseases are associated with increased levels of CCL5 and inversely associated with breast cancer risk [32, 33]. All this evidence supports the role of plasma CCL5 in breast tumor carcinogenesis.

In addition, both plasma CCL19 and CCL19 gene expression in tumors was associated with increased survival of breast cancer in our study. As downregulation of the CCL19/CCR7 axis has been observed in metastatic breast cancer compared to the primary tumor[34] and CCL19 suppresses angiogenesis and proliferation of several other types of cancers [35, 36], it is biologically plausible that CCL19 inhibits breast cancer metastasis through similar AIM2 and Met/ERK/Elk-1/HIF-1*α*/VEGF-A pathways [37] and therefore increases patient survival.

Interestingly, we also found that many of the associations between chemokines and breast cancer risk or survival were more pronounced in luminal A-or ER-positive cancers, suggesting that the effect of chemokines might be related to estrogen-related carcinogenesis [38]. However, we still found a potential causal association between CCL16 and triple-negative breast cancer. Further study is therefore needed to confirm this association and test its potential role as a biomarker or treatment target for triple-negative breast cancer.

The strength of our study included the use of the most recent and largest summary statistics for breast cancer with subtype-specific information and a comprehensive dataset for plasma chemokines. Our study had several limitations. As no sex-specific GWAS for chemokines has been reported, we assumed that the sex difference in the chemokine GWAS was small and that the original GWAS had already adjusted for age, sex and principal components. The generalizability of the findings is also affected by the use of European ancestry, and the results should be replicated in other populations.

## Conclusion

In conclusion, we found that lower plasma CCL5 was causally associated with higher breast cancer risk, while plasma and tumor-derived CCL19 was associated with increased survival of breast cancer patients. Both findings were more pronounced in ER-positive patients, indicating the potential use of CCL5 and CCL19 as biomarkers or drug targets in future studies for breast cancer prevention and treatment.

## Data Availability

Summary statistics for pQTLs of plasma chemokines can be downloaded from the IEU open GWAS project (https://gwas.mrcieu.ac.uk/). Summary statistics for breast cancer risk and survival are available on the BCAC website. (http://bcac.ccge.medschl.cam.ac.uk/). Data from the UK Biobank (http://www.ukbiobank.ac.uk/) are available to all researchers upon making an application.

## ETHICS APPROVAL AND CONSENT TO PARTICIPATE

All studies used for the calculation of breast cancer GWAS were approved by the relevant research ethics committee, and all participants provided informed consent.

## AVAILABILITY OF DATA AND MATERIALS

Summary statistics for pQTLs of plasma chemokines can be downloaded from the IEU open GWAS project (https://gwas.mrcieu.ac.uk/). Summary statistics for breast cancer risk and survival are available on the BCAC website. (http://bcac.ccge.medschl.cam.ac.uk/). Data from the UK Biobank (http://www.ukbiobank.ac.uk/) are available to all researchers upon making an application. Part of this research was conducted using the UK Biobank Resource under Application 61083.

## AUTHOR CONTRIBUTIONS

XY and HY had full access to all data and take responsibility for the integrity of the data and the accuracy of the analysis.

Study concept and design: HY

Acquisition, analysis, and interpretation of data: XY

Drafting of the manuscript: XY, YZ

Critical revision of the manuscript for important intellectual content: HY, XY, YL, FF

Statistical analysis: XY, YZ

Funding: HY.

## Supplementary Tables

sTable 1. List of the SNPs used as the genetic instrument for two-sample MR analysis

sTable 2. Causal association between plasma chemokines and overall and subtypes of breast cancer using different methods of two-sample MR analysis

sTable 3. Causal association between plasma chemokines and overall and subtypes of breast cancer survival using different methods of two-sample MR analysis

sTable 4. The potential causal association between plasma chemokine and major risk factors of breast cancer

